# LFSPROShiny: an interactive R/Shiny app for prediction and visualization of cancer risks in families with deleterious germline *TP53* mutations

**DOI:** 10.1101/2023.08.11.23293956

**Authors:** Nam H Nguyen, Elissa B Dodd-Eaton, Gang Peng, Jessica L. Corredor, Wenwei Jiao, Jacynda Woodman-Ross, Banu K. Arun, Wenyi Wang

**Author notes:** Corresponding author: Wenyi Wang, PhD; Department of Bioinformatics and Computational Biology, The University of Texas MD Anderson Cancer Center, 1400 Pressler Street, Houston, TX 77030;.

## Abstract

**Purpose:** LFSPRO is an R library that implements risk prediction models for Li-Fraumeni syndrome (LFS), a genetic disorder characterized by deleterious germline mutations in the *TP53* gene. To facilitate the use of these models in clinics, we developed LFSPROShiny, an interactive R/Shiny interface of LFSPRO that allows genetic counselors (GCs) to perform risk predictions without any programming components, and further visualize the risk profiles of their patients to aid the decision-making process.

**Methods:** LFSPROShiny implements two models that have been validated on multiple LFS patient cohorts: a competing-risk model that predicts cancer-specific risks for the first primary, and a recurrent-event model that predicts the risk of a second primary tumor. Starting with a visualization template, we keep regular contact with GCs, who ran LFSPROShiny in their counseling sessions, to collect feedback and discuss potential improvement. Upon receiving the family history as input, LFSPROShiny renders the family into a pedigree, and displays the risk estimates of the family members in a tabular format. The software offers interactive overlaid side-by-side bar charts for visualization of the patients’ cancer risks relative to the general population.

**Results:** We walk through a detailed example to illustrate how GCs can run LFSPROShiny in clinics, from data preparation to downstream analyses and interpretation of results with an emphasis on the utilities that LFSPROShiny provides to aid decision making.

**Conclusion:** Since Dec 2021, we have applied LFSPROShiny to over 100 families from counseling sessions at MD Anderson Cancer Center. Our study suggests that software tools with easy-to-use interfaces are crucial for the dissemination of risk prediction models in clinical settings, hence serving as a guideline for future development of similar models.

## Background

Germline mutations in cancer susceptibility genes can significantly increase the likelihood of developing certain cancer types during a person’s lifetime^1^. Risk prediction models built on family cancer history and Mendelian transmission to assess such increased cancer risks have been widely developed^2–6^. A prominent example is Li-Fraumeni syndrome (LFS)^7^, a rare autosomal-dominant hereditary disorder characterized by deleterious germline mutation in the *TP53* tumor suppressor gene^7^ that significantly increases the risks of a spectrum of cancer diagnoses, most notably breast cancer, soft-tissue sarcoma and osteosarcoma^7,8^. Hence, when assessing cancer risks for LFS patients, there exist additional challenges to quantify the risk of each cancer type separately. To address this issue, Shin et al (2019)^9^ proposed a Bayesian semi-parametric model that estimates cancer-specific age-at-onset penetrances for the first primary cancer. Since multiple primary cancers are particularly prevalent among LFS patients, Shin et al (2020)^10^ developed a complementary model that estimates penetrances beyond the first primary cancer, but does not differentiate between the cancer types. We will refer to the two models as the cancer-specific (CS) model and the multiple primary cancer (MPC) model. The penetrance estimates from the two models can be utilized for personalized risk predictions on unseen patients given their family history. The risk predictions have been validated on independent patient cohorts^11,12^, and have been shown to outperform the currently used clinical criteria when predicting the carrier status of *TP53* mutations^13^, including the Classic^14^ and the Chompret criteria^15,16^. We implemented the risk prediction models in a publicly available R package, called LFSPRO.

Conversations with families whose family history is suggestive of LFS regarding clinical decisions such as genetic testing and cancer screening, have been challenging. This is partly due to the fact that clinicians and genetic counselors (GCs) could only provide general, as compared to personalized, lifetime risks associated with LFS^17^. The good predictive performance of LFSPRO supports its potential utility to assist medical professionals with risk communication and decision-making in real-world clinical settings, hence providing a plausible solution to this bottleneck. For this purpose, it is highly desirable to have an interactive computer program that can be readily executed by the end users without dealing with the programming components. In this paper, we present LFSPROShiny, an R/Shiny implementation of LFSPRO that provides GCs and clinicians with an easy-to-use interface, comprehensive information on patients’ risk landscapes, and effective visualizations.

## Methods

### Overview of LFSPRO

LFSPRO is an R package that implements risk predictions based on penetrance estimates from two statistical models^9,10^. Both models were trained and validated on independent LFS family datasets^11,12^. The CS model^9^ was motivated by the fact that LFS patients are at increased risks of different cancer types^7,8^, which can be regarded as competing risks. Let *K* be the number of competing outcomes. The CS model computed the cancer-specific age-at-onset penetrance, denoted by 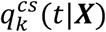. This is defined as the probability of developing cancer of type *k* by age *t* given the patient’s covariates ***X***. We considered four outcomes (i.e. *K* = 4): (i) breast cancer, (ii) sarcoma, (iii) all other cancer types combined, and (iv) competing mortality. The MPC model was motivated by the frequent cases of MPC among LFS patients. The model pooled all cancer types together, but extended beyond the first primary cancer. Let *L* be the number of cancer occurrences. The MPC model estimated the age-at-onset penetrance 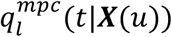, which is defined as the probability of developing the *l*th primary cancer by age *t* given the patient’s covariates and previous cancer occurrence at time *u*. We restricted our attention to the first and second primary cancers (i.e., *L* = 2).

LFSPRO utilizes the CS model to predict cancer-specific risks for the first primary cancer. While useful for drawing generic epidemiological insights, the penetrance estimates cannot immediately be used for risk prediction in clinics because they do not take into account the patient’s current age, denoted by *a*_*p*_. Let *T* denote the age at diagnosis of the first primary. The probability of developing the *k*th cancer type within the next *m* years, given that the patient has not developed cancer up to age *a*_*p*_, is given by

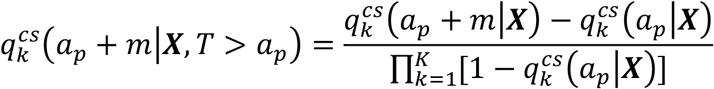

Due to the strong detrimental effect of *TP53* mutations, one of the most important covariates is the patient’s genotype (*G* = 1 for mutation carrier, *G* = 0 for wildtype). Most patients, however, have not undergone genetic testing (i.e., *G* is unknown). LFSPRO estimates the posterior probability that a patient carries deleterious germline *TP53* mutation given his or her family history. Let *G*_0_ be the genotype of the patient and ***H*** = {*H*_1_ *…, H*_*n*_} be the history of the *n* family members. Following our previous study^13^, we set the prevalence of *TP53* mutations in the general population, denoted by *P*[*G*_0_ = 1], to be 0.0006. LFSPRO computes the posterior probabilities *P*[*G*_0_ = *g*|***H***], *g* ∈ {0,1}, by assuming Mendel transmission. We refer the readers to **Supplementary Materials** for the detailed computations. The risk of developing the *k*th cancer type within the next *m* years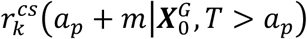, is then given by a weighted sum

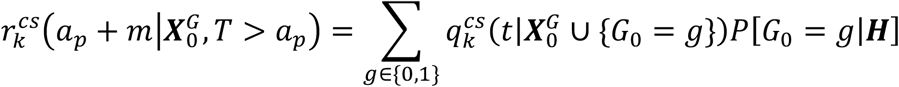

where 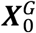, denotes the covariates without genotype.

LFSPRO utilizes the MPC model to predict the risk of a first primary cancer for patients who have not had cancer, and the risk of a second primary cancer for those who have had one primary cancer. Let *T*_*l*_ denote the age at diagnosis of the *l*th primary cancer. The probability of developing the *l*th primary cancer in the next *m* years given the patient’s current age *a*_*p*_ and the previous cancer occurrence at *t*_*l*−1_ is given by

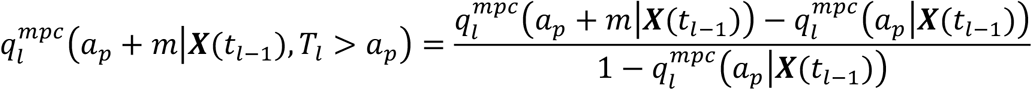

LFSPRO then follows similar, albeit not identical, calculations to compute the risk estimates for patients with unknown genotypes. Currently, LFSPRO is configured to report the 5-year, 10-year and 15-year risks.

### Development of LFSPROShiny

Prior programming knowledge in the R statistical software (v4.2.2; R Core Team 2022) is required to install and run LFSPRO. In clinics, it is highly desirable for GCs or clinicians to have a software tool that can be run without programming components. Furthermore, LFSPRO offers numerical output without any visualization capability. Not only does effective visualization help GCs to quickly understand the risk landscapes of their patients and make informed decisions regarding cancer screening and monitoring, but it also aids communication of cancer risks with the patients to justify such vital decisions. These shortcomings motivated us to develop LFSPROShiny, an R/Shiny implementation of LFSPRO that offers the end users an easy-to-use and interactive interface.

The development of LFSPROShiny was an iterative process. The first version offered a basic graphical user interface (GUI) and functionality. The development team was in regular contact with the GCs at MD Anderson Cancer Center (MDACC) to continually improve the application. Although most of the discussions took place in weekly meetings, the GCs also reached out to us when they encountered errors or unexpected results when running LFSPROShiny. Some changes were made to fix technical errors that resulted in incorrect risk predictions, and to improve the application’s robustness to deal with a variety of datasets that did not conform to the expected input data (e.g., family members with missing ages at cancer diagnosis or ages at last contact). Usually, such changes had to be implemented in the underlying LFSPRO package. Nevertheless, most of the changes were made to add useful functionalities to the application and to improve visualization. Overlaid bar charts were added for quick comparison between a patient’s predicted risks and the general population. The plots were further made interactive for easy downloading and to better show the numerical values of the bars.

The implementation of LFSPROShiny consists of: (1) a ui.R file that determines the overall GUI of the application, including the widgets (e.g., file input, radio button) and the output components (e.g., plot, table), (2) a server.R file that processes the input and displays the output on the application interface, and (3) a functions.R script, which contains user-defined functions that are invoked by server.R, in addition to the dependencies, to aid processing of the input. The dependencies include shiny, shinyjs, shinyalert, kinship2, DT, data.table, reshape2, ggplot2, ggsci and LFSPRO.

### User interface of LFSPROShiny

LFSPROShiny provides a GUI that requires no programming knowledge (**Figure 1**). Following the instructions in **Figure 1A**, GCs supply two input datasets that are formatted as .csv files (**Figure 1B**). The first is a family dataset that provides information such as age-at-last-contact, gender and *TP53* mutation status (if known) of the family members, each of whom is uniquely identified by an identification number, or ID. For each family member, the dataset contains the IDs of his or her parents, which can be used by LFSPROShiny to construct a pedigree plot based on the inferred familial relationship (**Figure 1C**). The second dataset describes the cancer occurrences, including the cancer types and ages at diagnosis. The cancer types must be coded according to the LFSPRO cancer spectrum. **Table 1** provides descriptions of the columns in the input datasets. Examples are provided in the **Results** section.

**Table 1:**
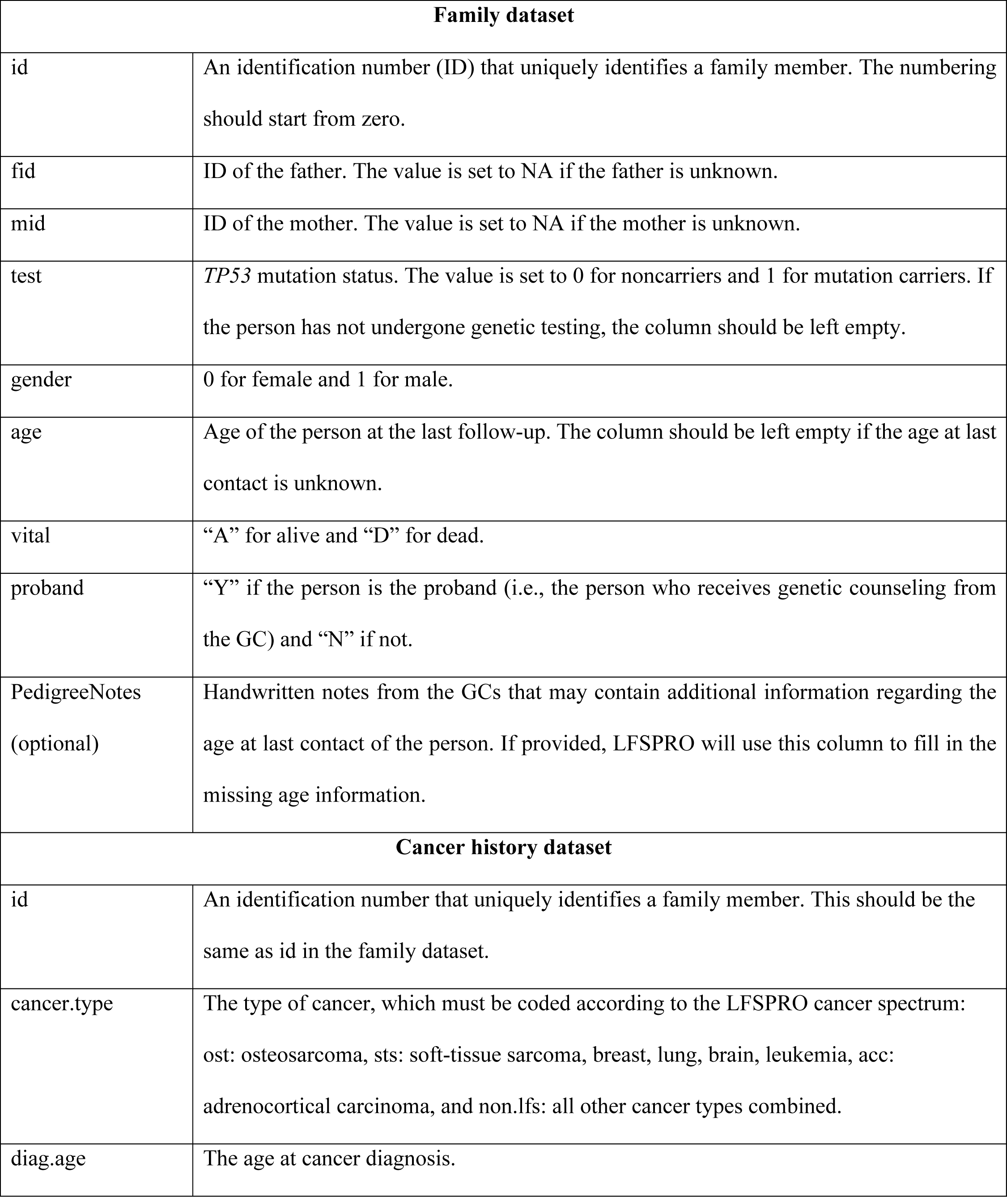
Column-by-column descriptions of the input family dataset and cancer history dataset.

| **Family dataset** |  |
| --- | --- |
| id | An identification number (ID) that uniquely identifies a family member. The numbering should start from zero. |
| fid | ID of the father. The value is set to NA if the father is unknown. |
| mid | ID of the mother. The value is set to NA if the mother is unknown. |
| test | *TP53* mutation status. The value is set to 0 for noncarriers and 1 for mutation carriers. If the person has not undergone genetic testing, the column should be left empty. |
| gender | 0 for female and 1 for male. |
| age | Age of the person at the last follow-up. The column should be left empty if the age at last contact is unknown. |
| vital | “A” for alive and “D” for dead. |
| proband | “Y” if the person is the proband (i.e., the person who receives genetic counseling from the GC) and “N” if not. |
| PedigreeNotes<br>(optional) | Handwritten notes from the GCs that may contain additional information regarding the age at last contact of the person. If provided, LFSPRO will use this column to fill in the missing age information. |
| **Cancer history dataset** |  |
| id | An identification number that uniquely identifies a family member. This should be the same as id in the family dataset. |
| cancer.type | The type of cancer, which must be coded according to the LFSPRO cancer spectrum:<br>ost: osteosarcoma, sts: soft-tissue sarcoma, breast, lung, brain, leukemia, acc:<br>adrenocortical carcinoma, and non.lfs: all other cancer types combined. |
| diag.age | The age at cancer diagnosis. |

**Figure 1:**
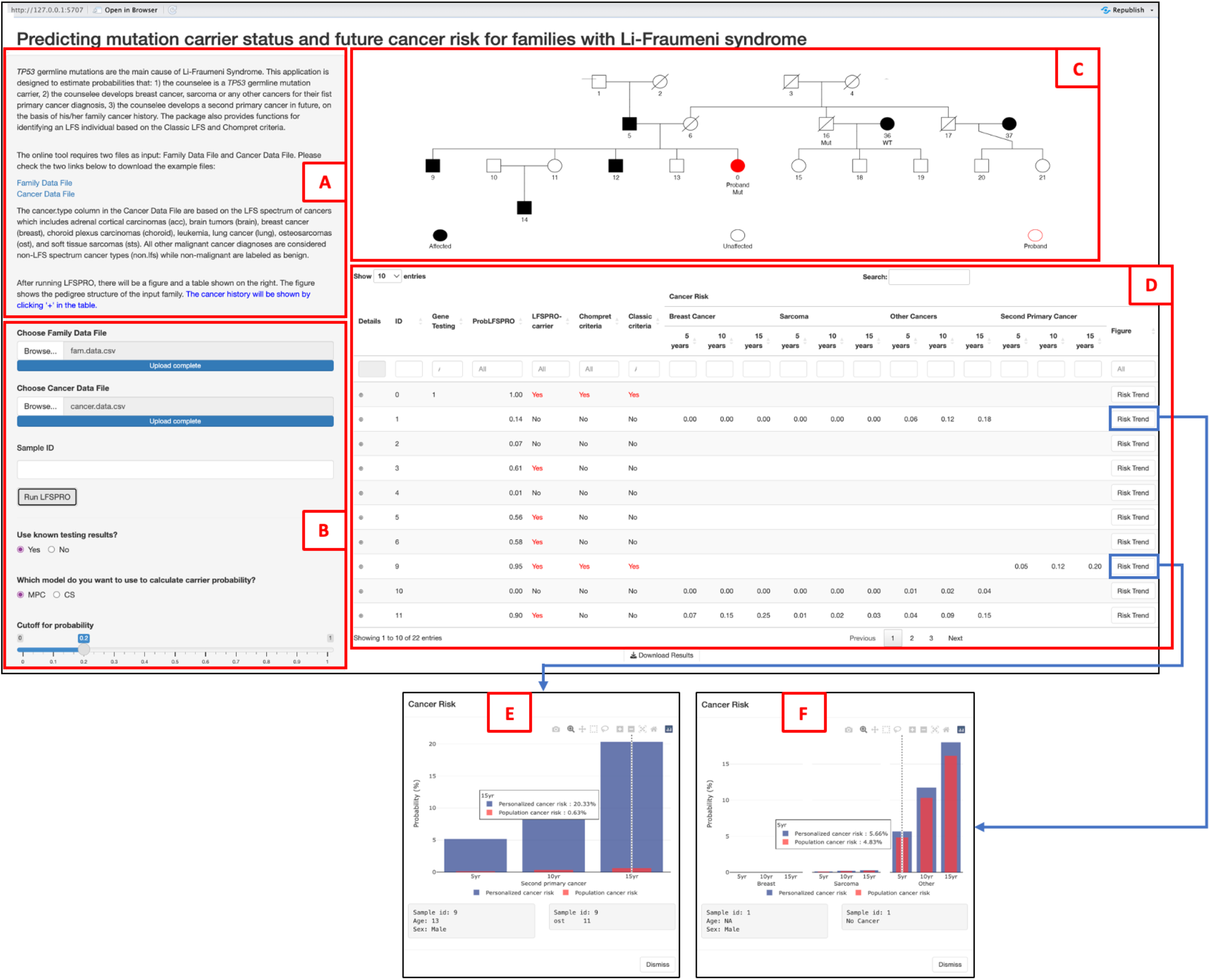
(A) Instructions for using LFSPROShiny; (B) Control panel for uploading the input datasets, running LFSPROShiny, and customizing the output; (C) Pedigree plot describing the relationship among family members, as constructed from the input family dataset; (D) Output describing the risk profile of each family member, including the probabilities of being TP53 mutation carrier from the CS and MPC models; (E) Cancer-specific risk prediction from the CS model for a patient with no cancer history; (F) Risk of a second primary cancer for a patient with one primary cancer occurrence in the past.

The output is displayed in a tabular format (**Figure 1D**), in which each record corresponds to a patient in the family. The main results of LFSPRO are the carrier probabilities for patients that have not undergone genetic testing, as well as the cancer risk predictions (as detailed below). By default, LFSPROShiny incorporates the *TP53* mutation status of family members who have been tested, but also provides an option on the left panel (**Figure 1B**) to disable this information. The MPC model is preselected to compute the carrier probabilities, but LFSPROShiny has an option for the users to choose the CS model (**Figure 1B**). A patient is predicted to be a germline *TP53* mutation carrier if the output probability is greater than or equal to 0.2. This decision threshold was shown to achieve good trade-offs between sensitivity and specificity on multiple LFS datasets^13^. If necessary, GCs can set their preferred cutoff values (**Figure 1B**). Prediction results from the Classic^14^ and Chompret^15,16^ criteria, which are the current standards for detection of *TP53* mutation carriers, are also provided.

For patients without cancer, LFSPROShiny invokes the CS model to provide separate probabilities for sarcoma, breast cancer, and all other cancer types combined, in the next 5, 10, and 15 years. For those who have already had one primary cancer, LFSPROShiny invokes the MPC model to calculate the probability of a second primary malignancy. LFSPROShiny plots the patient’s predicted risks, along with a hypothetical noncarrier of the same age and gender, in an overlaid interactive bar chart (**Figures 1E** and **1F**). Since LFS is a rare disorder^18^, it is reasonable to assume that the risks of noncarriers are representative of the general population.

### Deployment of LFSPROShiny

LFSPROShiny is supported across all major operating systems (Windows, macOS, and Linux). The source code is available on GitHub (https://github.com/wwylab/LFSPRO-ShinyApp). In the repository, the users can also find links to the underlying LFSPRO package. For public use, we host the live app on Shinyapps.io, which provides a convenient way to access LFSPROShiny on the web (https://namhnguyen.shinyapps.io/lfspro-shinyapp-master/). For clinical use at MDACC, LFSPROShiny is deployed on a virtual server, which is part of a VMware cluster. The virtual server has 16 processor cores and 64GB of memory, and operates on the Red Hat Enterprise Linux 7 operating system. This internal hosting structure ensures sufficient security controls for protected health information, and allows for immediate input of patient data without access issues. Since Dec 2021, we have applied LFSPROShiny to over 100 families from counseling sessions at MDACC, as GCs run LFSPROShiny on the family history data right after finishing the counseling session .

## Results

We provide a walk-through of how GCs can run LFSPROShiny in clinics. Due to the confidentiality of patient data, we create hypothetical family and cancer history datasets. **Figure 2 (left)** shows an example family dataset, and **Figure 2 (right)** shows the cancer history of this family. GCs may include additional columns that contain their notes regarding the patients’ ages at death. These columns are based on the handwritten notes that they take when additional information becomes available after the counseling sessions. LFSPROShiny automatically extracts information from the notes to complete the missing ages at last contact. For example, the person with ID 2 is estimated to have died at age 65.

**Figure 2:**
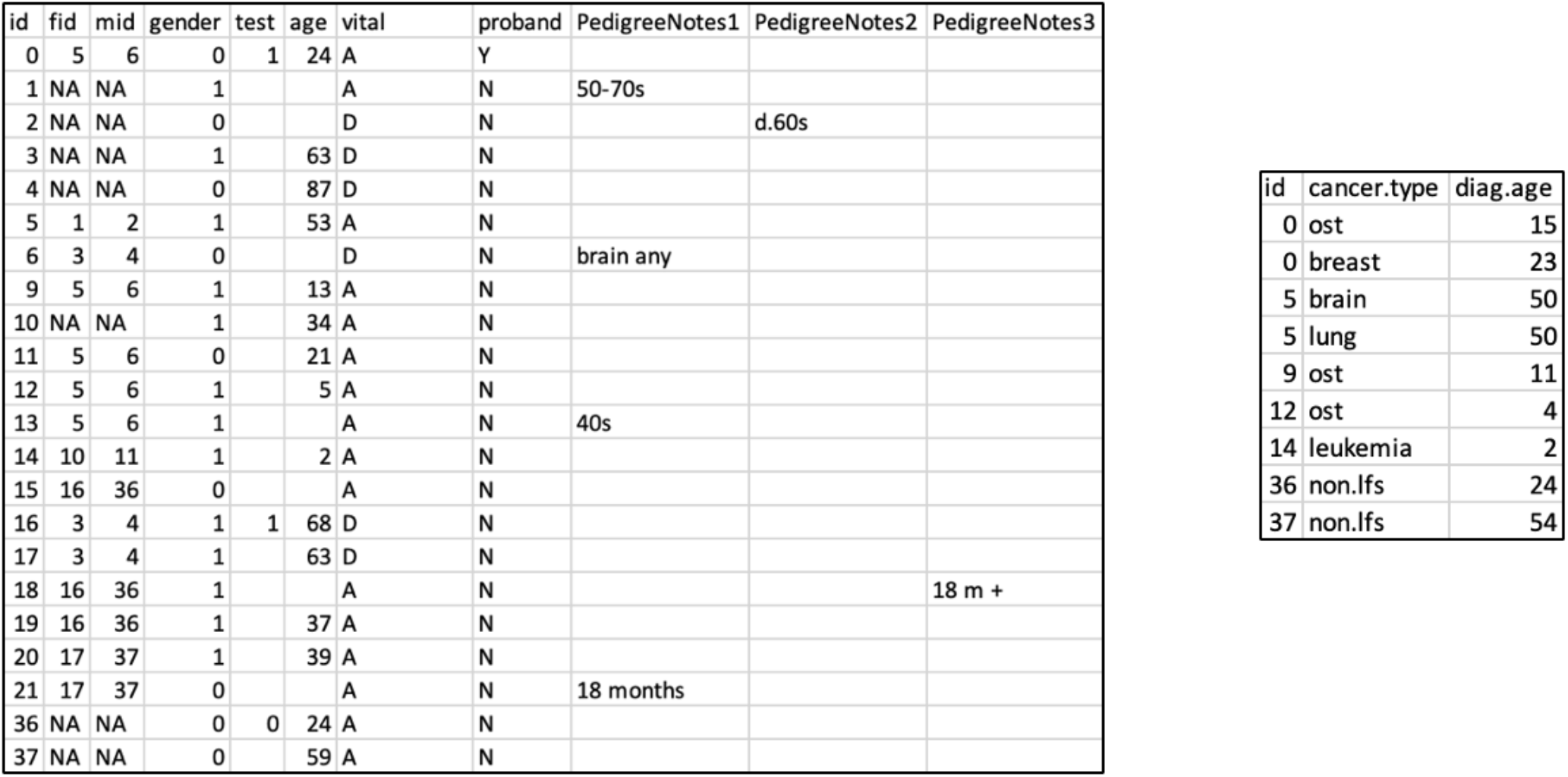
Family dataset with the GCs’ notes in the last three columns (left) and cancer history dataset that describes the cancer occurrences among family members (right).

These datasets, once formatted as csv files, can be uploaded to LFSPROShiny for downstream analyses. In **Supplementary Materials**, we provide addition examples to show how LFSPROShiny robustly handles incomplete datasets. LFSPROShiny infers the relationships among the family members based on their IDs, and graphically displays the family as a pedigree tree (**Figure 3**).

**Figure 3:**
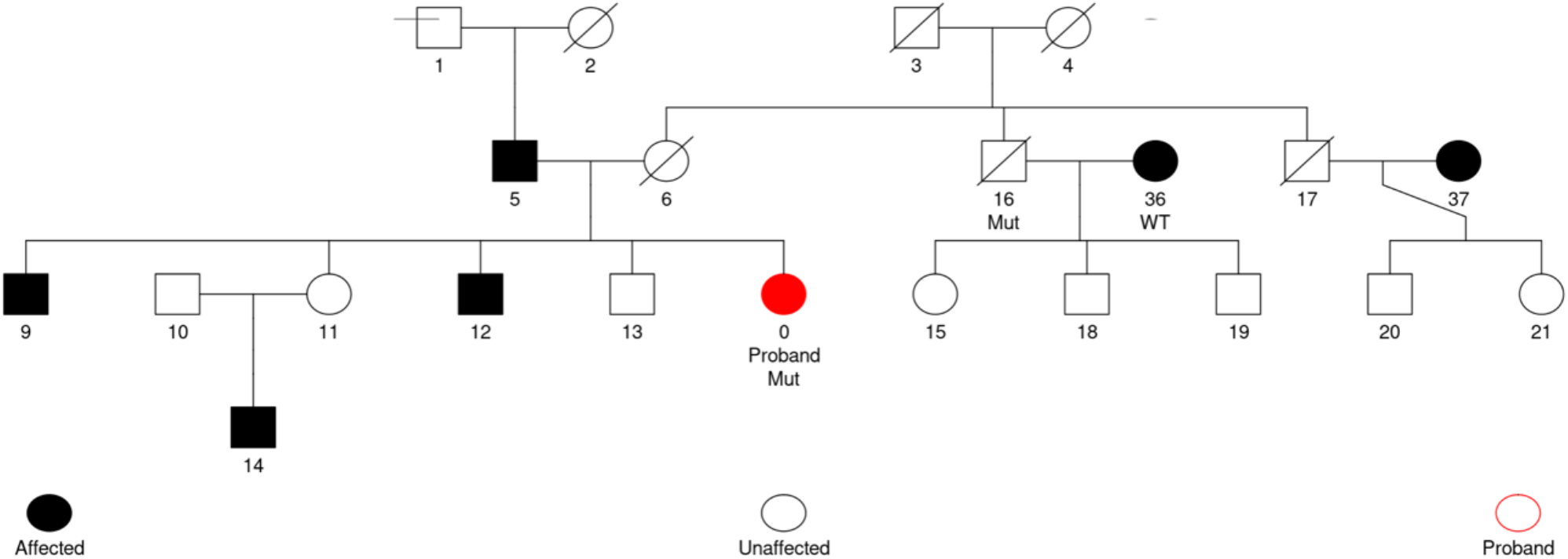
Pedigree tree that describes the relationship among family members. By convention, square denotes male and circle denotes female, and deceased family members are crossed with a diagonal. The proband is displayed in red. Individuals with at least one cancer occurrence (i.e., affected) is color-filled. Family members, who have undergone genetic testing, are annotated with their test results, with Mut denoting mutation carriers and WT denoting wildtypes.

One of the most important outputs is the probability of carrying a deleterious germline *TP53* mutation for an untested family member. GCs can use these numbers to identify at-risk individuals and potentially recommend genetic testing for them. **Figure 4** shows the mutation probabilities from the CS and MPC models. The tab for each family member can be expanded to show basic information and cancer history. Using the MPC model, the predicted mutation probability for ID 5 is 0.56, whereas the CS model gives a near-zero probability. This observation can be explained by the characteristics of the two models. The CS model focuses on the first primary cancer only: it was not until 50 years old that the person developed brain cancer, which, although part of the LFS spectrum, is much less prevalent than breast cancer and sarcoma among LFS patients. Hence, there is little evidence to predict that the person has *TP53* mutation from the perspective of the CS model. On the other hand, the MPC model, which does not differentiate between cancer types, considers only the fact that the person has had two primary cancers, leading to a much higher probability. The CS model might have overlooked the second primary tumor and, consequently, underestimated the carrier probability. Had the first primary cancer be early-onset osteosarcoma, the CS model might have been the better choice since it would have perceived this cancer occurrence as a strong signal for LFS. Currently, we provide both models in LFSPROShiny for GCs to use them on a case-by-case basis. We also discuss a potential solution in the **Conclusion** section. From **Figure 3**, the proband, who has been tested positive, is an offspring of IDs 5 and 6. Given that the proband has two siblings with osteosarcoma at very young ages (IDs 9 and 12), it is highly likely that her *TP53* mutation is not de novo and that one of the parents is also a mutation carrier. Hence, in the CS model, the near-zero probability for ID 5 is accompanied by a probability that is close to 1 for ID 6. The Chompret criteria does not take into account this information and predicts wildtype for both IDs 5 and 6.

**Figure 4:**
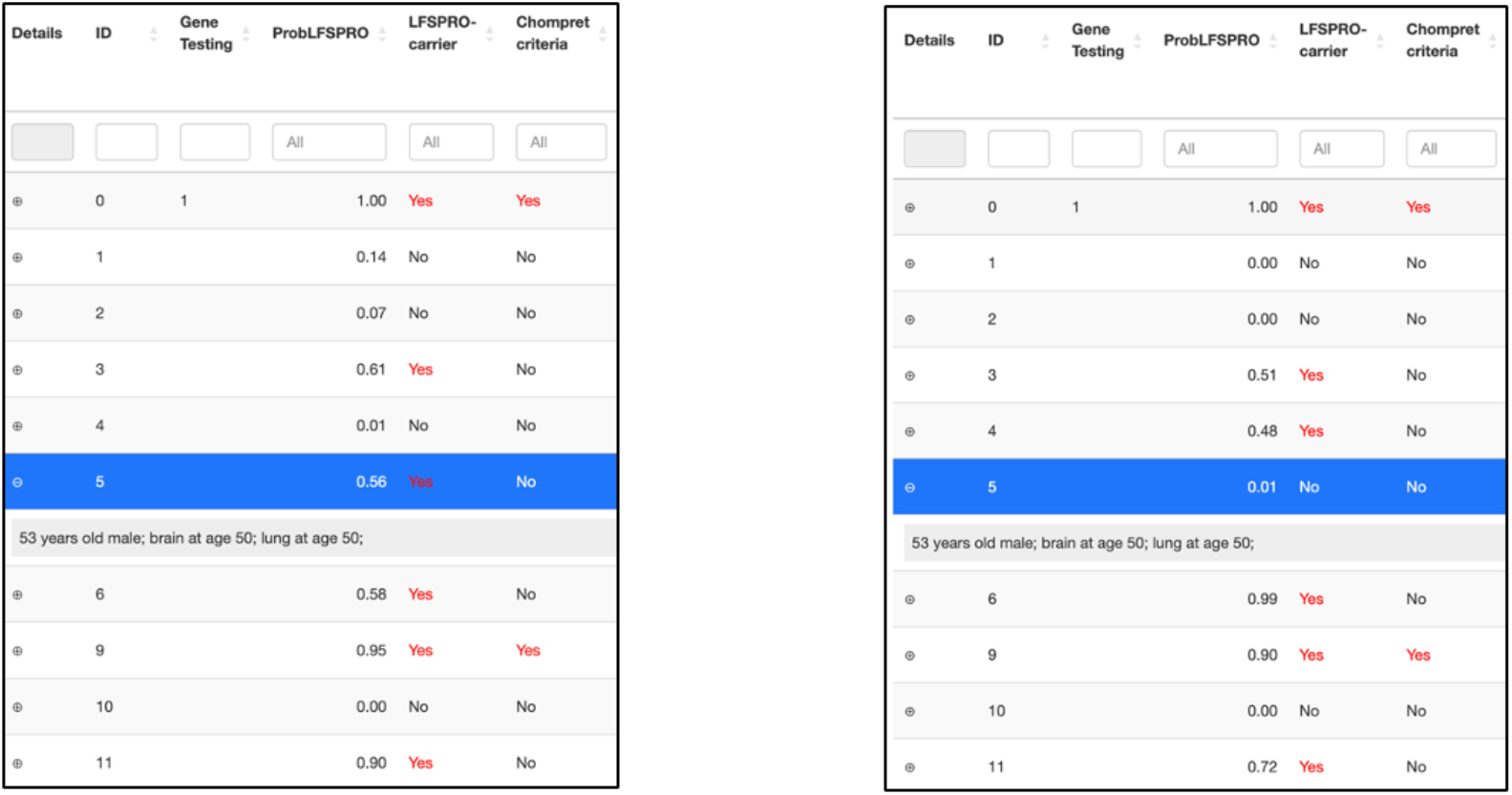
TP53 mutation probabilities (ProbLFSPRO) produced by the MPC model (left) and the CS model (right). The binary decision (LFSPRO-carrier) is determined by the default cutoff probability of 0.2. The Shiny app allows users to modify the cutoff probability. Prediction from the Chompret criteria, which is part of the current NCCN guideline is also provided.

LFSPROShiny provides interactive overlaid bar charts for GCs to visualize the cancer risks of their patients relative to the general population. For ID 9, who has already had osteosarcoma at age 11, LFSPROShiny invokes the MPC model to compute the risk of a second primary tumor (**Figure 5, top left**). This individual has a very high mutation carrier probability (**Figure 4**), thus the predicted cancer risks (blue) are well above the general population (red). ID 1 has not had cancer, so LFSPROShiny invokes the CS model to compute the risks of breast cancer, sarcoma, and all other cancer types combined, for the first primary cancer (**Figure 5, top right**). This person is predicted at near-zero probability to carry a mutation (**Figure 4**), hence the predicted cancer risks (blue) are only slightly higher than the general population (red). The overall risk of developing a first primary tumor can then be computed by summing up the cancer-specific risks. Risk prediction is not available for ID 4, who already died at age 87 (**Figure 5, bottom left**). Currently, LFSPROShiny does not offer the capability to predict cancer-specific risks beyond the first primary, hence risk prediction is not available for ID 5, who has already had brain and lung cancers (**Figure 5, bottom right**).

**Figure 5:**
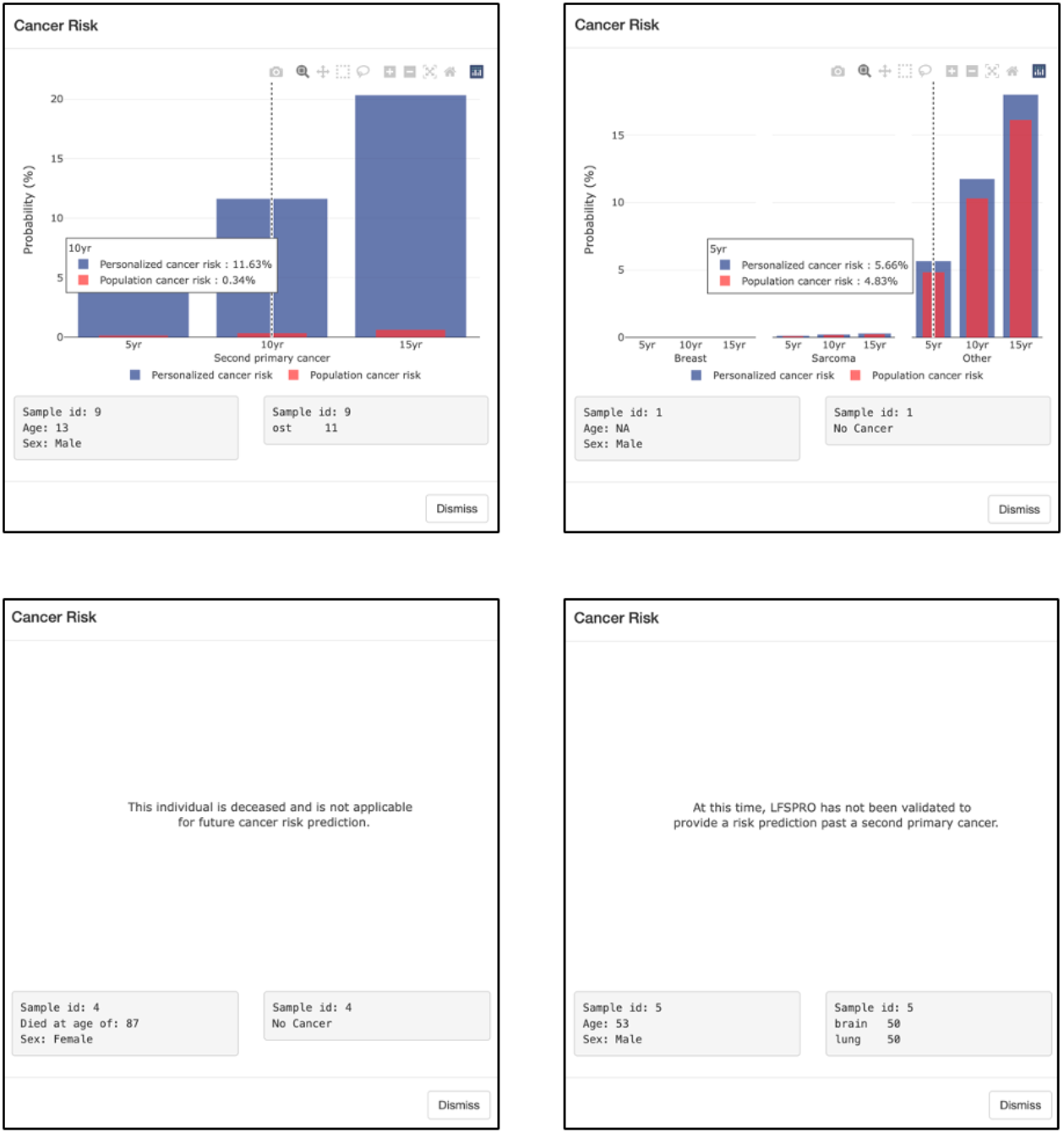
LFSPROShiny predicts the risks of a second primary tumor for individuals with one cancer (top left), and predicts cancer-specific risks (breast cancer, sarcoma, and all other cancer types combined) for those without cancer history (top right). The personalized risks are shown in blue, while the risks in the general population are shown in red. The predicted risks are provided for the next 5, 10 and 15 years. Risk predictions are not available for deceased individuals (bottom left), and for those who have had more than one cancer occurrence (bottom right).

## Conclusion

In this paper, we present LFSPROShiny, a GUI software application that is built upon the previously validated LFSPRO R package to provide a much-needed interface for medical professionals to calculate, visualize and assess a patient’s personalized mutation carrier probability and cancer risks based on their family history. At-risk patients can be advised accordingly to pursue genetic testing and, if appropriate, undergo the current LFS cancer screening protocols for timely detection of cancer development. Providing patients with personalized risk assessment can help patients better understand their diagnosis and may motivate them to follow the current annual cancer screening regime, which remains an intensive and sometimes overwhelming process. The application requires no programming knowledge, is supported across multiple platforms, and produces the output in seconds. Thus, GCs can access the application on their own computers to assist real-time decision making in their counseling sessions. Our study suggests that software tools with GUI and minimum requirements of programming knowledge can improve and expedite the clinical utility of personalized cancer risk predictions.

By continually increasing the user group of GCs, LFSPROShiny will be expanded to include additional functionalities. Currently, the software can only be used to predict cancer-specific risks of the first primary cancer for patients who have never had cancer, and the risks of a second primary malignancy for those who have had one primary cancer. Risk predictions for cancer survivors are becoming increasingly important. A recent study reported that there were over 18 million living cancer survivors in the United States on January 1, 2022^19^. This number is projected to exceed 20 million by 2026 and 26 million by 2040^20^. Numerous studies have confirmed the dependence of the subsequent primary tumors on the type and timing of the previous occurrences^21,22^. Nevertheless, the current clinical management of cancer survivors is largely indifferent from at-risk individuals, thus not accounting for these relationships. To address the needs in this growing population, we will expand LFSPROShiny to include a unified model that predicts cancer-specific risks beyond the first primary cancer, recently developed by Nguyen et al (2023)^23^, after it is further validated on multiple independent patient cohorts. We will also update the existing CS and MPC models as needed. Given the generality of these frameworks, they can be retrained on more complex family datasets to include additional competing risks or extend the risk predictions to the third primary cancer. Additional features, such as the type of treatment for the first primary cancer, can be included to improve the models’ performance^24–26^. Finally, the visualization components of LFSPROShiny will continue to be updated to further advance effective risk communication in the new generation of genetic and clinical counseling.

## Data Availability

The source code of LFSPROShiny and example datasets are publicly available on GitHub (https://github.com/wwylab/LFSPRO-ShinyApp). The live app is hosted on Shinyapps.io (https://namhnguyen.shinyapps.io/lfspro-shinyapp-master/). The underlying LFSPRO package can be found on GitHub (https://github.com/wwylab/LFSPRO).

https://namhnguyen.shinyapps.io/lfspro-shinyapp-master

https://github.com/wwylab/LFSPRO

## Acknowledgement

The authors thank Dr. Seung Jun Shin and Jingxiao Chen for their contributions to LFSPRO.

