## Supplementary materials for "LFSPROShiny: an interactive R/Shiny app for prediction and visualization of cancer risks in families with deleterious germline *TP53* mutations"

#### A. Computation of carrier probability

Let $G_0$ be the genotype of a patient, who has not undergone genetic testing, and $\boldsymbol{H} = \{H_1, \ldots, H_n\}$ be the cancer history of his or her family members. We follow our previous study[1] and set the prevalence of pathogenic *TP53* mutations in the general population to be 0.0006. From the Hardy-Weinberg equilibrium, it follows that the prevalence of wildtype ($G_0 = 0$), heterozygous mutation ($G_0 = 1$) and homozygous mutation ($G_0 = 2$) are 0.9988, 0.0005996 and 3.6e-07, respectively. LFSPRO computes the posterior probabilities $P[G_0 = g|\boldsymbol{H}]$, $g \in \{0,1,2\}$, via the Bayes rule as follows[2]

$$P[G_0|\boldsymbol{H}] = \frac{P[G_0]P[\boldsymbol{H}|G_0]}{\sum_{G_0} P[G_0]P[\boldsymbol{H}|G_0]}$$

where

$$P[\boldsymbol{H}|G_0] = \sum_{G_1,\ldots,G_n} P[\boldsymbol{H}|G_0, G_1, \ldots, G_n]P[G_1, \ldots, G_n|G_0]$$

$$= \sum_{G_1,\ldots,G_n} \left[\prod_{i=0}^{n} P[H_i|G_i]\right] P[G_1, \ldots, G_n|G_0]$$

It is reasonable to assume that the family members are conditionally independent given the genotypes, hence the conditional family-wise likelihood, $P[\boldsymbol{H}|G_0, G_1, \ldots, G_n]$, factorizes into a product of the conditional individual likelihoods, $P[H_i|G_i]$. Calculations of $P[H_i|G_i]$ are different for the CS and MPC models. We refer the readers to the original research papers[3,4] for more details of the two models. Assuming Mendelian transmission, we can compute the probability $P[G_1, \ldots, G_n|G_0]$, but the computation can be very complex for large families. Thus, we employ the Elston-Stewart peeling algorithm[5] to calculate $P[\boldsymbol{H}|G_0]$ recursively in an efficient way while accounting for the pedigree structure of the family. To do this, we first pick an individual in the family to be the pivot. We then split the family into two disjoint subsets: (i) the anterior consists of family members that are related to the pivot through his or her parents, and (ii) the posterior consists of those that are related to the pivot through his spouse or offsprings. Conditional on the pivot's genotype, the anterior and posterior are independent, hence

$$P[\boldsymbol{H}|G_0] = \sum_{G_p} P\left[\boldsymbol{H}_p^-|G_p, G_0\right]P\left[H_p|G_p\right]P\left[\boldsymbol{H}_p^+|G_p, G_0\right]$$

where $G_p$ denotes the genotype of the pivot, $\boldsymbol{H}_p^-$ denotes the cancer history of the anterior part, and $\boldsymbol{H}_p^+$ denotes the cancer history of the posterior part. $P\left[H_p|G_p\right]$ represents the likelihood contribution of the pivot, which can be computed from the CS or MPC model. The anterior and posterior probabilities are computed in the same way by randomly picking a pivot individual within each subset. The essence of the peeling algorithm is shown in **Supplementary Figure 1**.

**[Supplementary Figure 1]**

#### B. Handling of incomplete datasets

##### B.1. Parent ID is missing or not found

LFSPROShiny uses the peeling algorithm[5] to incorporate the pedigree structure into calculating *TP53* mutation probabilities. The algorithm requires that a person has both the father ID and the mother ID, or neither of them (i.e., the person is at the top of the pedigree tree in this case). Clinical data as encountered by the GCs in their counseling sessions, however, are not always complete. Sometimes, a family member has a missing father ID or mother ID, but not both. In this case, LFSPROShiny takes the following steps to automatically infer the missing parent to ensure the application's robustness:

1. LFSPROShiny first browses through the family to find family members who share the same non-missing parent ID as the person in question. If they exist and their other parent ID is available, then LFSPROShiny will complete the missing data with this parent ID for the person in question. If there are multiple parent IDs, then one of them will be random selected to fill in the missing data.
2. If the step 1 is unable to resolve the issue, LFSPROShiny creates a dummy parent for the family member.

Additionally, a family member may have a parent ID that is not found within the input family dataset due to mistakes in data entry. In this case, this parent ID is treated as missing, and LFSPROShiny follows the steps above to fill in such missing data.

**Supplementary Figure 2** shows an example family, in which a family member has a missing parent ID, and the missingness can be resolved by step 1 (i.e., without creating a dummy parent).

**[Supplementary Figure 2]**

**Supplementary Figure 3** shows another example, in which the missing parent ID cannot be resolved by step 1. Hence, LFSPROShiny creates a dummy parent, which appears in the pedigree tree (**Supplementary Figure 4**).

**[Supplementary Figure 3]**

**[Supplementary Figure 4]**

**B.2. Uninformative family**

In real clinical settings, the input family and cancer history datasets that the GCs obtain from their patients may not contain much information. In some cases, this is due to a lack of follow-up time so that the current family remains almost cancer-free. While LFSPRO has been shown to accurately predict *TP53* mutations[1] and cancer risks[6,7] without any genetic testing results in the family, the predictions can be unreliable under extreme lack of cancer history. For this reason, a family is considered uninformative if (i) there is at most one cancer occurrence among the family members and (ii) there is at most one family member who has been tested. **Supplementary Figure 5** shows an example of an uninformative family. In this case, LFSPROShiny produces a warning message for the GCs to be aware of the lack of information (**Supplementary Figure 6**). This rule will be updated with more data in future studies.

**[Supplementary Figure 5]**

**[Supplementary Figure 6]**

### Supplementary figures

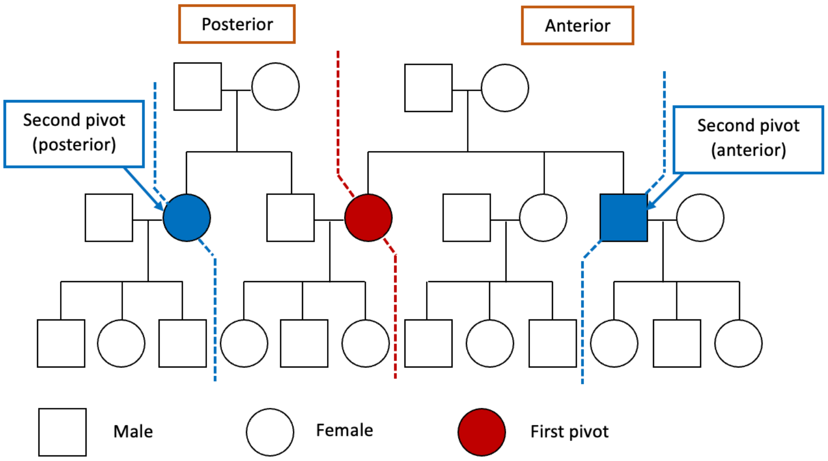


*Supplementary figure 1: The peeling algorithm recursively splits the pedigree into two disjoint subsets based on a randomly selected individual (pivot). The two subsets are independent given the pivot's genotype.*

| id | fid | mid | gender | test | age | vital | proband | PedigreeNotes1 | PedigreeNotes2 | PedigreeNotes3 |
|---|---|---|---|---|---|---|---|---|---|---|
| 0 | 5 | 6 | 0 | 1 | 24 | A | Y | | | |
| 1 | NA | NA | 1 | | | A | N | 50-70s | | |
| 2 | NA | NA | 0 | | | D | N | | d.60s | |
| 3 | NA | NA | 1 | | 63 | D | N | | | |
| 4 | NA | NA | 0 | | 87 | D | N | | | |
| 5 | 1 | 2 | 1 | | 53 | A | N | | | |
| 6 | 3 | 4 | 0 | | | D | N | brain any | | |
| 9 | 5 | 6 | 1 | | 13 | A | N | | | |
| 10 | NA | NA | 1 | | 34 | A | N | | | |
| 11 | 5 | 6 | 0 | | 21 | A | N | | | |
| 12 | 5 | 6 | 1 | | 5 | A | N | | | |
| 13 | 5 | 6 | 1 | | | A | N | 40s | | |
| 14 | 10 | 11 | 1 | | 2 | A | N | | | |
| 15 | 16 | 36 | 0 | | | A | N | | | |
| 16 | 3 | 4 | 1 | 1 | 68 | D | N | | | |
| 17 | 3 | 4 | 1 | | 63 | D | N | | | |
| 18 | 16 | 36 | 1 | | | A | N | | | 18 m + |
| 19 | 16 | 36 | 1 | | 37 | A | N | | | |
| 20 | 17 | 37 | 1 | | 39 | A | N | | | |
| 21 | 17 | NA | 0 | | | A | N | 18 months | | |
| 36 | NA | NA | 0 | 0 | 24 | A | N | | | |
| 37 | NA | NA | 0 | | 59 | A | N | | | |

*Supplementary figure 2: A family dataset, in which the family member with ID 21 has a missing mother ID. LFSPROShiny manages to locate the person with ID 20, who shares the same father (ID 17) but has a non-missing mother (ID 37). Then, LFSPROShiny uses ID 37 as the mother of ID 21 in the subsequent calculations.*

| id | fid | mid | gender | test | age | vital | proband | PedigreeNotes1 | PedigreeNotes2 | PedigreeNotes3 |
|---|---|---|---|---|---|---|---|---|---|---|
| 0 | 5 | 6 | 0 | 1 | 24 | A | Y | | | |
| 1 | NA | NA | 1 | | | A | N | 50-70s | | |
| 2 | NA | NA | 0 | | | D | N | | d.60s | |
| 3 | NA | NA | 1 | | 63 | D | N | | | |
| 4 | NA | NA | 0 | | 87 | D | N | | | |
| 5 | 1 | 2 | 1 | | 53 | A | N | | | |
| 6 | 3 | 4 | 0 | | | D | N | brain any | | |
| 9 | 5 | 6 | 1 | | 13 | A | N | | | |
| 10 | NA | NA | 1 | | 34 | A | N | | | |
| 11 | 5 | 6 | 0 | | 21 | A | N | | | |
| 12 | 5 | 6 | 1 | | 5 | A | N | | | |
| 13 | 5 | 6 | 1 | | | A | N | 40s | | |
| 14 | 10 | 11 | 1 | | 2 | A | N | | | |
| 15 | 16 | 36 | 0 | | | A | N | | | |
| 16 | 3 | 4 | 1 | 1 | 68 | D | N | | | |
| 17 | 3 | 4 | 1 | | 63 | D | N | | | |
| 18 | 16 | 36 | 1 | | | A | N | | | 18 m + |
| 19 | 16 | 36 | 1 | | 37 | A | N | | | |
| 20 | 17 | NA | 1 | | 39 | A | N | | | |
| 21 | 17 | NA | 0 | | | A | N | 18 months | | |
| 36 | NA | NA | 0 | 0 | 24 | A | N | | | |
| 37 | NA | NA | 0 | | 59 | A | N | | | |

*Supplementary figure 3: A family dataset, in which the family members with ID 20 and ID 21 both has a missing mother ID. Since LFSPROShiny cannot locate another person who shares the same father, it will automatically create a dummy parent with ID 38 and assign her to be their mothers.*

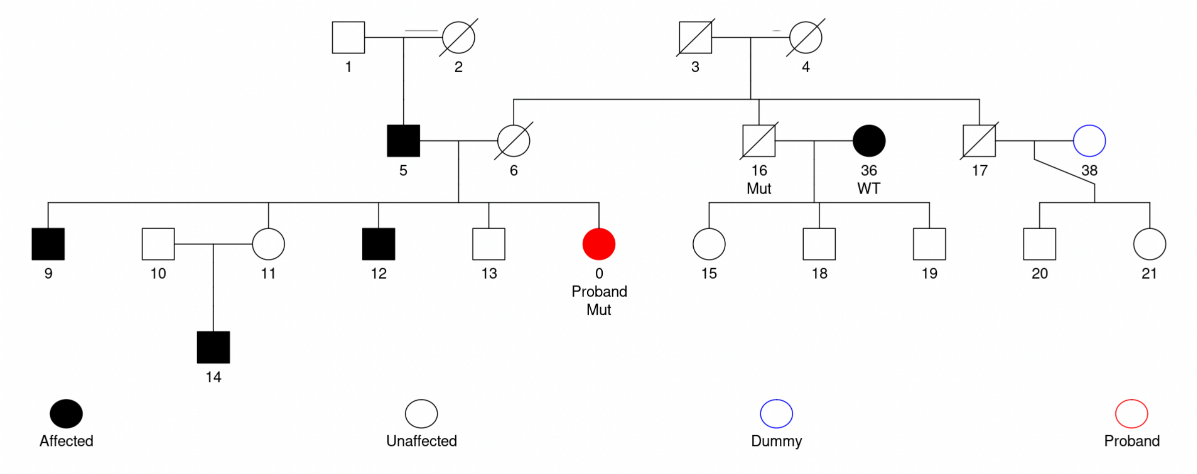


*Supplementary Figure 4: A pedigree tree that shows the dummy parent in blue (ID 38).*

| id | fid | mid | gender | test | age | vital | proband | PedigreeNotes1 | PedigreeNotes2 | PedigreeNotes3 |
|---|---|---|---|---|---|---|---|---|---|---|
| 0 | 5 | 6 | 0 | 1 | 24 | A | Y | | | |
| 1 | NA | NA | 1 | | | A | N | 50-70s | | |
| 2 | NA | NA | 0 | | | D | N | | d.60s | |
| 3 | NA | NA | 1 | | 63 | D | N | | | |
| 4 | NA | NA | 0 | | 87 | D | N | | | |
| 5 | 1 | 2 | 1 | | 53 | A | N | | | |
| 6 | 3 | 4 | 0 | | | D | N | brain any | | |
| 9 | 5 | 6 | 1 | | 13 | A | N | | | |
| 10 | NA | NA | 1 | | 34 | A | N | | | |
| 11 | 5 | 6 | 0 | | 21 | A | N | | | |
| 12 | 5 | 6 | 1 | | 5 | A | N | | | |
| 13 | 5 | 6 | 1 | | | A | N | 40s | | |
| 14 | 10 | 11 | 1 | | 2 | A | N | | | |
| 15 | 16 | 36 | 0 | | | A | N | | | |
| 16 | 3 | 4 | 1 | | 68 | D | N | | | |
| 17 | 3 | 4 | 1 | | 63 | D | N | | | |
| 18 | 16 | 36 | 1 | | | A | N | | | 18 m + |
| 19 | 16 | 36 | 1 | | 37 | A | N | | | |
| 20 | 17 | 37 | 1 | | 39 | A | N | | | |
| 21 | 17 | 37 | 0 | | | A | N | 18 months | | |
| 36 | NA | NA | 0 | | 24 | A | N | | | |
| 37 | NA | NA | 0 | | 59 | A | N | | | |

| id | cancer.type | diag.age |
|---|---|---|
| 0 | ost | 15 |

*Supplementary figure 5: Example of an uninformative family. Only the proband (ID 0) had a cancer occurrence (osteosarcoma at age 15), and he is the only family member who has undergone genetic testing.*

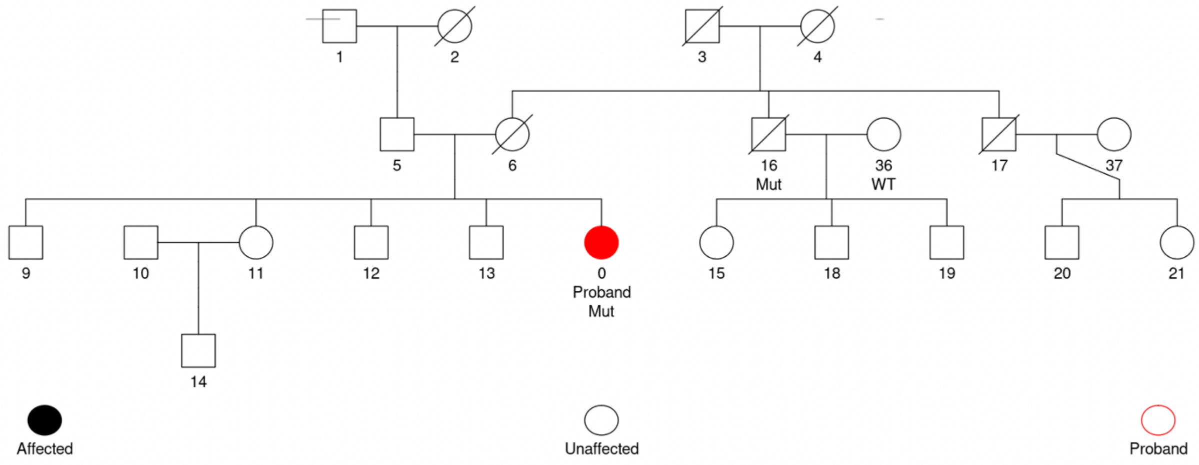


Warning: This family has limited information; The predicted risks can be unreliable.

*Supplementary figure 6: A pedigree tree with a warning message for limited information.*
